# Ecological and Immune Pressures Shape Outcomes of Precision Phage Therapy in Advanced Cystic Fibrosis Lung Disease

**DOI:** 10.1101/2025.05.22.25327316

**Authors:** Tiffany Luong, Lukeman Kharrat, Kevin Champagne-Jorgensen, Jennifer A. Melendez, David Pride, Douglas J. Conrad, Dwayne R. Roach

## Abstract

Chronic *Pseudomonas aeruginosa* infection remains a defining and increasingly drug-resistant complication in cystic fibrosis (CF). Here, we report phage-antibiotic co-therapy in an elderly individual with CF and a multidrug-resistant pulmonary exacerbation. Treatment was safe, well-tolerated, and associated with rapid clinical improvement, including enhanced lung function, reduced obstruction, and a 100-fold decrease in bacterial burden. Multi-omic profiling revealed *in situ* phage replication, transient suppression of non-mucoid *P. aeruginosa*, and virome restructuring. Bacterial isolates evolved resistance with associated fitness costs, while the host developed phage-specific neutralizing antibodies that limited systemic activity. Despite this, one phage persisted in the lung beyond therapy. These findings offer *in vivo* insight into phage–host– microbiome dynamics during treatment and underscore the importance of integrating microbial ecology and immune profiling into phage therapy design. This case highlights the potential for phage therapy in late-stage CF and the value of personalized, immune-informed strategies for managing chronic infection.

## INTRODUCTION

*Pseudomonas aeruginosa* is a predominant pathogen responsible for chronic respiratory infections in people with cystic fibrosis (pwCF), with its ability to rapidly adapt to environments, develop antibiotic resistance, and persist in the hostile CF lung environment making it a significant therapeutic challenge ^1^. The bacterium thrives in the thick, viscous mucus of the CF lung, where its virulent mechanisms, including biofilm formation and genomic plasticity, complicate treatment regimens and hinder pathogen eradication ^2,3^. As *P. aeruginosa* increasingly evolves into multidrug-resistant (MDR) and extensively drug-resistant (XDR) strains, the efficacy of standard antibiotics continues to diminish, accelerating disease progression and driving a rise in the frequency of pulmonary exacerbations ^3,4^. These exacerbations, characterized by sudden declines in lung function, accelerate the deterioration of pulmonary health and severely impair the quality of life of CF patients ^1^. Although cystic fibrosis transmembrane conductance regulator (CFTR) modulators offer clinical benefits by partially correcting the underlying ion transport defect ^5^, they do not fully address the persistent *P. aeruginosa* infections that continue to drive much of the disease’s morbidity ^6^.

Consequently, there is a critical need for innovative therapeutic approaches, such as bacteriophage (phage) therapy, that can effectively target MDR *P. aeruginosa* strains and offer a promising adjunct or alternative to conventional antibiotics, potentially improving both the management of pulmonary exacerbations and long-term outcomes for pwCF. Phage therapy uses viruses that specifically target and kill bacterial cells through the process of infecting a bacterial host, hijacking its cellular machinery to replicate, and ultimately causing cell lysis. Phages offer distinct advantages over traditional antibiotics, particularly in their different mechanisms of action, ability to combat biofilm-associated bacteria, and infection of MDR and XDR strains. More than 6000 patients have received phage therapy as single patient compassionate use or in clinical trials since its advent ^7–9^. However, phage therapy has not yet received regulatory approval in most countries ^10^.

A growing body of evidence supports the potential of phage therapy as a viable treatment option for refractory bacterial infections. Several studies have demonstrated the safety and efficacy of phages in preclinical and clinical settings, with phage therapy showing promise in improving pulmonary function in pwCF ^7,8^. Additionally, phage-antibiotic combination therapies have shown enhanced therapeutic effects, overcoming the limitations of antibiotics alone ^11,12^. However, the clinical application of phage therapy in CF, and beyond, remains limited by unknown factors such as phage selection criteria, optimal dosing, pharmacokinetics, phage resistance, and host immune response to the biologic. Despite these challenges, the safety and potential efficacy of phage therapy in CF treatment underscores the need for more comprehensive studies. Furthermore, the potential of phages to complement antibiotic therapy, particularly for XDR strains, has yet to be fully explored.

In this study, we investigate the combined use of phage therapy and ciprofloxacin to treat severe pulmonary exacerbation caused by *P. aeruginosa* in an elderly individual with CF (Figure 1a). We evaluate therapeutic impact through changes in lung function, obstruction clearance, and patient-reported outcomes. In parallel, we examine bacterial genomic and phenotypic shifts, microbiome and virome dynamics, and the development of anti-phage immunity. This case provides a rare opportunity to explore how phage therapy performs in late-stage disease and offers insights for optimizing treatment strategies as MDR pathogens become increasingly prevalent in CF.

**Figure 1:**
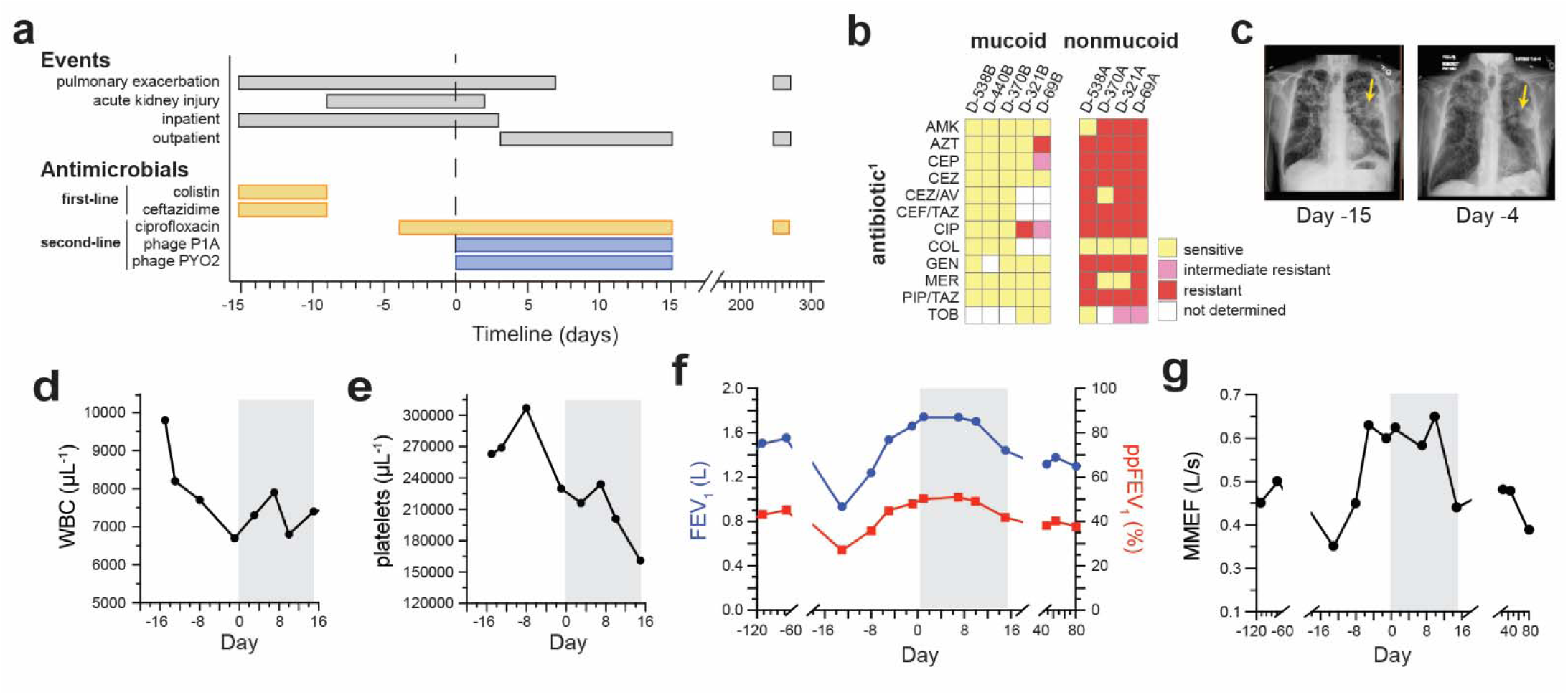
Antimicrobial treatment outcomes for a complex, acute *P. aeruginosa* pulmonary exacerbation in person with cystic fibrosis (pwCF). (a) Hospitalization event and antimicrobial treatment timeline of the patient with CF. Gray bars represent the duration of pulmonary exacerbation and acute kidney injury episodes, with inpatient and outpatient treatment designations indicated. Colored bars show the duration of antibiotic treatments (colistin, ceftazidime, ciprofloxacin; yellow) and phage therapies (P1A, PYO2; blue). Both antibiotics and phages were administered intravenously (i.v.). Timeline numbering is relative to initiating IV phages, indicated at the vertical dashed line (day 0). **(b)** Heatmap comparing the minimum inhibitory concentration (MIC) of patient sputa bacterial isolates to antibiotics (Table S2). Isolates were assessed by colony phenotype with mucoid (left) isolates being largely susceptible and non-mucoid (right) isolates exhibiting extensive resistance to antibiotics. ^1^Antibiotic abbreviations: amikacin (AMK), aztreonam (AZT), cefepime (CEP), ceftazidime (CEZ), ceftazidime/avibactam (CEZ/AV), ceftolozane/tazobactam (CEF/TAZ), ciprofloxacin (CIP), colistin (COL), gentamicin (GEN), meropenem (MER), piperacillin/tazobactam (PIP/TAZ), and tobramycin (TOB). **(c)** Posterior-to-anterior chest radiographs (x-rays) obtained at hospitalization for exacerbation (day −15, left) and shortly before initiation of the co-treatment (day −4, right) both demonstrating left upper lobe opacity (yellow arrow). The density of the opacity decreases but does not resolve between the two timepoints, indicating the persistence of the infiltrate. **(d-g)** Line graphs of patient clinical outcomes with the co-treatment period combining ciprofloxacin and phages highlighted in gray. **(d)** White blood cell (WBC) counts (cells·µL^-1^) and **(e)** platelet counts (cells·µL^-1^) measured from whole blood collection. **(f)** Spirometry measure of pulmonary change in patient forced expiratory volume (FEV_1_, blue) shown alongside values normalized for age, weight, and sex as percent predicted FEV_1_ (ppFEV_1_, red). **(g)** Calculation of mid-maximal expiratory flow (MMEF) rate (L/s) from spirometry.

## RESULTS

### Last resort antibiotics were safely substituted with a combination of antibiotic and phage

A 75–80-year-old male with CF (genotype c.3849+10kbC>T and c.2012delT) was hospitalized with a severe pulmonary exacerbation caused by a multidrug-resistant (MDR) *Pseudomonas aeruginosa* infection (Figure 1b) localized to the left upper lobe (Figure 1c). Given his advanced age, comorbidities, and longstanding chronic infection, his initial prognosis was poor (Table S1). In the year leading up to this event, sputum cultures consistently revealed two distinct *P. aeruginosa* phenotypes: a mucoid, pan-sensitive population and a non-mucoid population resistant to most antibiotic classes except polymyxins (Figure 1b, Table S2). The patient was started on low dose intravenous (IV) colistin (1 mg/kg every 12 hours) and ceftazidime (25 mg/kg every 8 hours), in combination with supportive therapies including inhaled hypertonic saline, DNase, bronchodilators, and supplemental oxygen (Figure 1a). Initial symptom improvement was interrupted by the development of colistin-induced acute kidney injury (AKI), necessitating cessation of antibiotic therapy after six days (Figure S2). At the time of colistin withdrawal, chest imaging continued to show a large, unresolved infiltrate (Figure 1c).

In response, second-line therapy with low dose ciprofloxacin (400 mg every 18 hours) and a two-phage cocktail was initiated to target both non-mucoid and mucoid variants of the infection (see Methods and Figure S2–S3). To reduce the risk of adverse effects or antimicrobial antagonism ^13^, phage and antibiotic administration were staggered by two days. A single test dose of phages (2 × 10 PFU in 1 mL dPBS) was administered with a saline flush to confirm tolerability. Following safety verification, phages were given every eight hours (2 × 10 PFU/mL) for the remainder of a standard 14-day antimicrobial course for pulmonary infections ^14^. The patient tolerated treatment well and stabilized after three days of inpatient phage therapy, allowing transition to outpatient care with continued monitoring. Phage dosing continued without interruption, adverse events, or the need for dose adjustments, indicating successful integration of phage therapy into the treatment regimen.

Upon admission, the patient had an elevated white blood cell (WBC) count (Figure 1d), indicative of an active immune response to the infection and stressed the infection’s severity. Treatment with colistin/ceftazidime and ciprofloxacin reduced WBC count to 6700 cells/µL over an initial 15 days, demonstrating the therapies’ ability to manage infection and modulate immune activity. Platelet counts, another biomarker of inflammation ^15^, were also elevated at hospitalization and peaked at 307,000 cells/µL after 4 days of colistin treatment (Figure 1e). The elevated platelet counts in conjunction with the rising creatinine levels (Figure S1) indicate systemic inflammation and the detrimental impact of colistin treatment on the patient’s kidney health ^16^. The discontinuation of colistin gradually brought platelet counts back down towards baseline (Figure 1d), signifying reduced inflammation.

Interestingly, phage therapy was associated with a renewed rise in both white blood cell (WBC) and platelet counts. WBCs increased to 7,300 cells/µL and remained elevated for seven days (Figure 1d), while platelet counts rose to 234,000 cells/µL (Figure 1e). This mirrors the immune response seen with certain antibiotics, such as tobramycin and beta-lactams, which elevate leukocyte levels via the release of bacterial components during lysis ^17^. We infer that phage-mediated bacterial lysis similarly triggered immune activation through the release of inflammatory bacterial products ^18^. Alternatively, the response may reflect direct immunogenicity of phage particles themselves, including structural proteins ^19^ and nucleic acids ^20^, or the early stages of antibody development ^21^. After therapy was discontinued, both WBC and platelet counts returned to near-baseline levels.

Spirometry provides critical insights into airway function and is essential for monitoring treatment response over time. At baseline, the patient showed moderate to severe airflow limitation, with a forced expiratory volume in one second (FEV_1_) of 1.49 L, or 43% of the predicted value based on age, weight, and sex (Figure 1f). Upon admission, FEV_1_ declined to 0.92 L, and maximal mid-expiratory flow (MMEF), a marker of small airway function ^22^, fell to 0.35 L/s (Figure 1g). Antibiotic therapy led to substantial improvement, with FEV_1_ rising to 1.66 L and MMEF to 0.6 L/s. Phage co-treatment was associated with an additional 0.08 L increase in FEV_1_, while MMEF showed a modest gain by day 10. Following treatment cessation, both FEV_1_ and MMEF declined slightly, to 96% and 92% of baseline values, respectively (Figure 1f–g). Importantly, no long-term decline in lung function or architecture was observed, with stable spirometry results maintained for at least 80 days after therapy.

### Phages boosted antibiotics to clear an airway obstruction

A marked reduction in forced vital capacity (FVC) is common in lung infections due to inflammation and fluid accumulation that limit exhalation ^23^. Upon admission, the patient’s FVC dropped to 0.92 L (37% predicted), indicating severe airway obstruction (Figure 2a). Both FEV1 and FVC were reduced, but the proportionally greater decline in FVC led to an elevated FEV_1_/FVC ratio (Figure 2b), consistent with impaired inhalation or reduced vital capacity. Antibiotic therapy increased FVC to 3.03 L (66% predicted), approaching baseline (Figure 2a). However, colistin-induced acute kidney injury (AKI) likely contributed to pulmonary edema, which corresponded with a reduced FEV_1_/FVC ratio on day 8 (Figure 2b), highlighting the interplay between renal and pulmonary function. Notably, the initiation of phage therapy was associated with a faster rate of FVC recovery, 0.18 L/day compared to 0.07 L/day with ciprofloxacin alone. After seven days of co-therapy, FVC rose to 6.75% above the patient’s baseline, suggesting that phages contributed additively to obstruction removal. However, FVC declined slightly by the end of treatment, indicating that antimicrobial effects were not fully sustained (Figure 2a).

**Figure 2:**
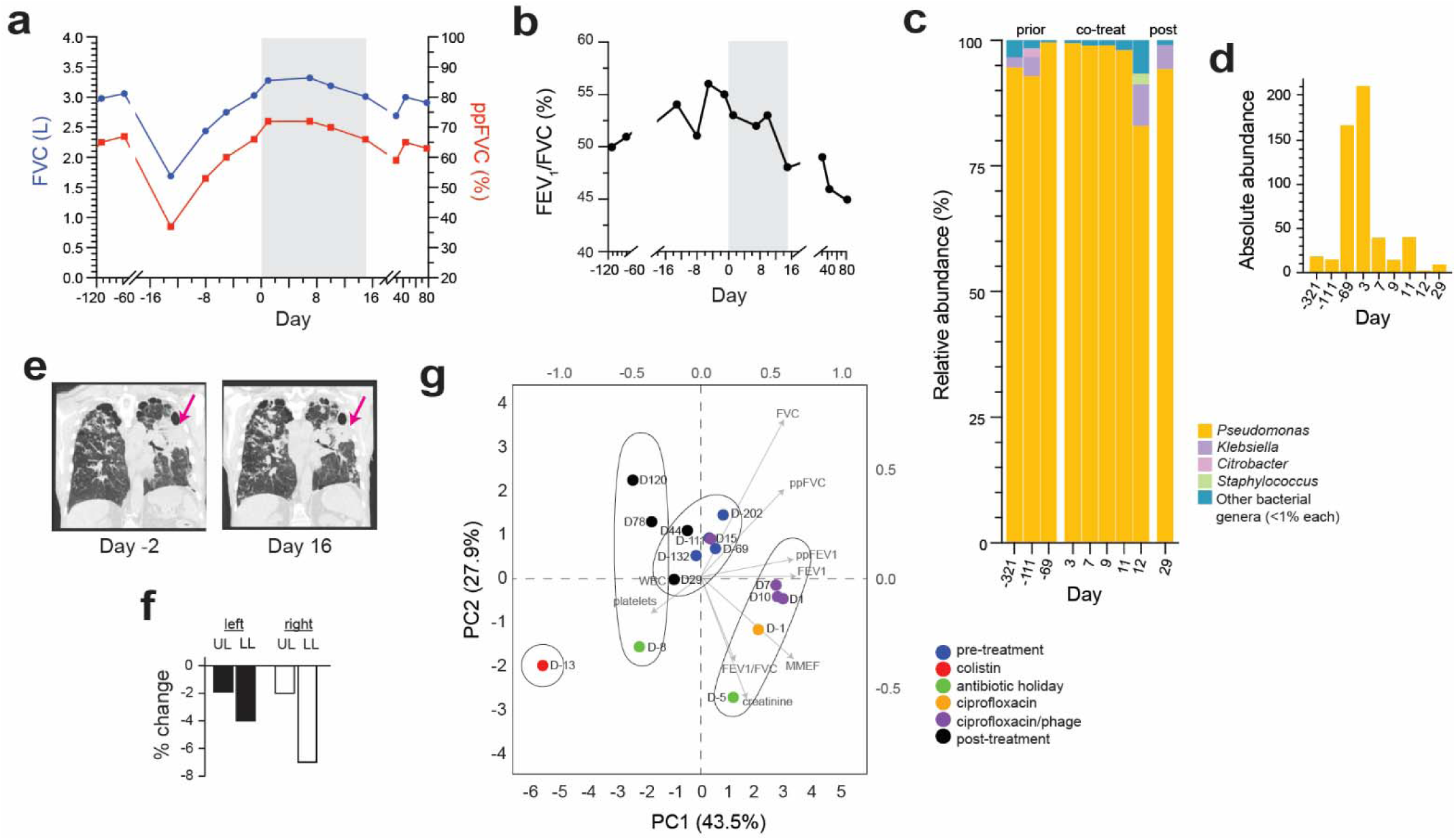
Co-treatment additively removes obstruction to improve recovery rate. **(a-b)** Line graphs of patient clinical outcomes with the co-treatment period combining ciprofloxacin and phages highlighted in gray. **(a)** Spirometric measurement of obstruction removal in forced vital capacity (FVC, blue) which is graphed alongside percent predicted FVC (ppFVC, red). **(b)** Ratio of FEV_1_/FVC expressed as percent change over time. **(c)** Stacked bar plots of relative abundance as fractions of total bacterial reads (%) preceding, during, and after co-therapy. Bars are colored by bacterial genera: *Pseudomonas* (yellow), *Klebsiella* (purple), *Citrobacter* (pink), *Staphylococcus* (green), and all other genera with fewer than 1% reads (blue). **(d)** Bar graph of absolute abundance of reads mapped to the *Pseudomonas* genus. Absolute abundance was calculated by normalizing bacterial reads against diploid human genomes within each sample. Whereas relative abundance shows >80% abundance across all treatment samples, the absolute abundance unravels patterns of *Pseudomonas* reduction which were overshadowed by microbiome dominance of the chronic infection. **(e)** Quantitative computed tomography (qCT) images of the chest in the axial lung window demonstrating progressive changes in pulmonary parenchymal abnormalities between day −2 (after 2 days of ciprofloxacin, left) and day 16 (1 day after co-treatment end, right). Decreased opacity of white regions indicates reduced left upper lobe inflammation (pink arrow). **(f)** Bar graph showing percent change, calculated from qCT scores in air trapping in each lung’s upper lobe (UL) and lower lobe (LL) 15 days after initiating phage treatment. **(g)** Principal component analysis (PCA) plot showing the distribution of dimensionally reduced patient sample data (spirometry, sera, and creatinine levels). Data points are labeled by date and colored according to the treatment: pre- treatment (blue), colistin (red), antibiotic holiday (green), ciprofloxacin (orange), ciprofloxacin/phage (purple), and post-treatment (black). Hierarchically determined clusters are circled. Vector loadings are shown in gray with arrows indicating each characteristic’s influence on the PCs.

Complementing spirometry findings, sputum metagenomics were used to investigate the microbial dynamics within the lungs. Sputa collected before exacerbation, during co-treatment, and post-treatment showed that the patient’s lungs were consistently dominated by *Pseudomonas* which accounted for >90% of bacterial reads in 8/9 sputum samples (Figure 2c, Table S3). Before exacerbation the patient’s lungs exhibited low bacterial diversity (5-7%) with only three non-*Pseudomonas* genera present in significant amounts: *Staphylococcus* and *Klebsiella*, both common CF pathogens ^24^, and *Citrobacter*, a transient colonizer. This limited diversity was eliminated before exacerbation when *Pseudomonas* reads increased to 99% on day −69. Co-treatment appeared to reduce *Pseudomonas* relative abundance to 83% after 11 days, which allowed other bacteria to expand in the lungs. *Klebsiella* increased from 3.8% to 8.4% while the other genre remained in low individual abundances (Table S3). Post-treatment the increased lung diversity was short-lived, and *Pseudomonas* reads increased to baseline, approx. 94%.

While high *Pseudomonas* relative abundance is typical in chronic CF lung infection, it can obscure comparisons of total bacterial burden across samples. To address this, we calculated absolute abundance by normalizing *Pseudomonas* reads to human reads, estimating bacterial cells per human cell ^25^. Between the days −111 and −69, *Pseudomonas* abundance increased 8-fold (15 to 167 cells per human cell), foreshadowing exacerbation onset (Figure 2d). Three days into phage therapy, levels remained elevated (211 cells per human cell), but co-treatment led to a 10-fold reduction by day 7, closely aligning with improved lung function (Figure 2a, 2d). Although bacterial levels remained near baseline during continued therapy, fluctuations in the second week may reflect evolving resistance (Figure 2d). Quantitative chest CT corroborated microbial clearance, showing reduced inflammation and improved airway patency (Figure 2e). A 3.26% average reduction in air trapping across lung lobes (Figure 2f, Table S4) further supports phage therapy’s role in obstruction relief.

We next performed principal component analysis (PCA) to assess the effects of antimicrobial treatments on patient health, using a dataset of serum, kidney, and lung function variables (Table S5). Lung health accounted for 71.4% of the variance, with FEV_1_ and FVC driving principal components 1 and 2 (PC1 and PC2), respectively (Figure 2g, Table S6). This reflects the central role of lung function in respiratory disease outcomes ^26^. Colistin/ceftazidime treatment shifted the patient’s profile along PC1, indicating improved airflow driven by increases in FEV_1_. Three distinct clusters emerged over time, corresponding to clinical changes. A brief “antibiotic holiday” (days -9 to -5) reversed this progress along PC2, reflecting reduced FVC and renewed obstruction. Phage therapy produced a sharper shift along PC2, suggesting a greater effect on FVC, consistent with obstruction clearance and bacterial load reduction. This aligns with preclinical findings that phages can rapidly reduce pulmonary infections ^12,27^. By treatment end, the patient returned to the baseline cluster (Figure 2g), indicating that therapeutic benefits were not sustained after all antimicrobials were stopped.

### Co-treatment was necessary to overcome phenotypic and genotypic diversity in CF

The patient’s mixed *Pseudomonas* infection, comprising pan-sensitive mucoid and MDR non-mucoid subpopulations, prompted a combined treatment approach with ciprofloxacin and phages. Efficacy was assessed by quantifying colony-forming units (CFUs) from sputum cultures. Ciprofloxacin alone produced a modest 0.5-log reduction in mucoid cells (2 × 10 CFU/mL/day) over four days, while fluoroquinolone-resistant non-mucoid cells increased by 2 logs (Figure 3a). With co-treatment, non-mucoid CFUs dropped by 3 logs within one day, indicating robust phage activity. Mucoid clearance also accelerated (3.2 × 10 CFU/mL/day), likely due to phage-induced biofilm disruption ^28^. However, by day 7, mucoid clearance plateaued, and phage efficacy against non-mucoid cells waned after day 3, followed by a 1-log rebound by day 11. By treatment end, total bacterial load declined from ∼10 to ∼10 CFU/mL, suggesting reversion to a lower-burden chronic state ^29^. Mucoid cells remained dominant (9.3 × 10 CFU/mL), while non-mucoid cells stabilized at ∼2 logs lower, representing a favorable shift toward a more antibiotic-susceptible profile.

**Figure 3:**
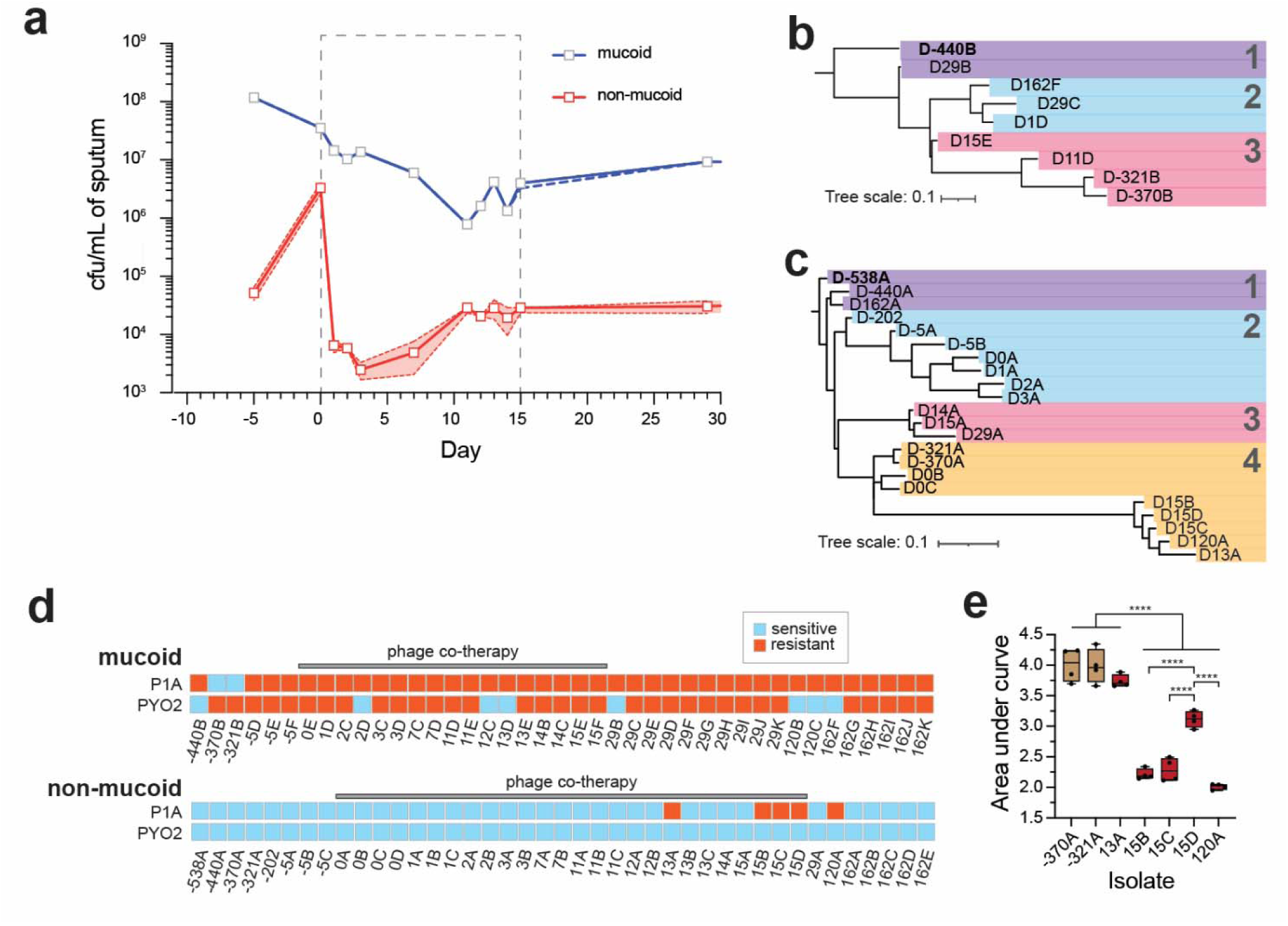
Additive co-treatment reduces target *Pseudomonas* density of both phenotypes. **(a)** Direct quantification of colony-forming units per mL (cfu/mL) of unenriched patient sputa on selective *Pseudomonas* media. Non-mucoid cfu counts are graphed in red and mucoid cfu counts in blue with a dashed box indicating the co-treatment period. Error bars are 95% confidence interval. **(b-c)** Maximum likelihood phylogenetic trees of clinical *P. aeruginosa* subpopulations made from accessory genome alignments generated with *Roary*. The tree was made using FastTree using a generalized time-reversible (GTR) model with 1000 iterations. Both trees have scale bars representing 0.1 nucleotide substitutions per site. Tree clusters are numbered and indicated by colored bars. **(b)** Tree rooted to the oldest mucoid isolate (D-440B, bolded) showing the relationship between 9 mucoid isolates sampled over time. **(c)** Tree rooted to the oldest non-mucoid clinical isolate (D-538A, bolded) showing the relationship between 22 non-mucoid isolates sampled over time. **(d)** Heatmap of clinical isolate susceptibilities to phages P1A and PYO2. Isolate morphology mucoid or non-mucoid were determined after 48h incubation on selective cetrimide agar. Each column represents a single isolate, labeled by sputa sample date. The gray bar above each set of isolates indicates the period (from days 0-15) of phage co-therapy. The ability to form plaques on a solid bacterial lawn at 10^8^ pfu/mL was considered sensitive (blue) and the lack of any clearance considered resistant (orange). **(e)** Boxplots comparing bacterial growth, measured by area under the curve (AUC), of clinical isolates grown for 24h in phage-free media. P1A-sensitive isolates are colored brown and P1A-resistant isolates are colored red. Boxplots show the median, interquartile range, and whiskers representing the data distribution. Asterisks indicate statistical significance ****p < 0.0001.

To assess *P. aeruginosa* evolution across treatment phases, we analyzed pangenome phylogenies of nine mucoid and twenty-two non-mucoid isolates. Core genome analysis revealed minimal variation within either subpopulation (Figure S4–S5), reflecting the conserved nature of genes essential for host survival ^30^. In contrast, mucoid isolates showed notable shifts in accessory genome content, genes found in fewer than 95% of strains and often linked to virulence and resistance ^31^ (Figure 3b). Cluster 1, spanning pre- and post- treatment isolates, appeared genetically stable, likely due to biofilm-associated persistence ^32^. Meanwhile, Cluster 3 isolates from pre-treatment and mid-treatment (days 11 and 15) showed increasing divergence, suggesting adaptation under treatment pressure. A new clade emerging post-phage therapy (days 1, 29, and 162) may reflect ciprofloxacin-driven evolution. These findings underscore the genetic adaptability and resilience of the mucoid phenotype during chronic infection.

Non-mucoid isolates exhibited pronounced accessory genome responses to phage therapy (Figure 3c). A stable subpopulation cluster with minimal divergence implied limited evolutionary aggressiveness, potentially facilitated by mixed population dynamics. Clade 2 isolates demonstrated early clonal expansion followed by significant divergence due to phage pressures, ultimately disappearing post-day 3. Clade 4 isolates showed resilience to antibiotics but substantial genetic divergence during late treatment (days 13, 15, and 120), indicative of evolution to a more phage resistant population. A new lineage emerged post-day 14, suggesting ongoing selective pressures or a novel environmental strain introduction. These rapid genomic adaptations emphasize the importance of continuous monitoring to optimize therapeutic strategies.

To link genomic variation to functional outcomes, we assessed phenotypic phage susceptibility across isolates collected during the 15-day co-treatment. The administered cocktail consisted of two lytic antipseudomonal phages (P1A and PYO2) that target distinct lipopolysaccharide (LPS) receptors (Figure S6–S7). Among mucoid isolates, phage susceptibility was limited: only 25% (10/41) were sensitive to either P1A or PYO2, but not both (Figure 3d). Sensitivity to P1A was rare before treatment (2/6 isolates) and absent thereafter, suggesting possible resistance emergence. This resistance may correlate with genomic divergence observed in clade 3 during the second treatment week (Figure 3b). In contrast, susceptibility to PYO2 remained stable throughout treatment, indicating resistance did not emerge. Overall, phage activity against the mucoid population was minimal, likely due to biofilm-associated protection ^3^.

Non-mucoid isolates were more susceptible, with 88% showing sensitivity to both phage types. However, P1A-resistant variants began appearing during the second treatment week, detectable on day 13. All five P1A-resistant isolates exhibited reddish-brown hyperpigmentation (Figure S8), a phenotype previously linked to *hmgA* mutations ^33^. Despite this emergence, P1A resistance remained low in frequency (15%), during treatment and 14% post-treatment, and did not dominate the lung population. This was likely due to an associated fitness cost as 4 out of 5 resistant isolates exhibited reduced growth rate (Figure 3e). These phenotypes aligned with the genetic divergence of clade 4 isolates (Figure 3c), supporting bacterial adaptation under strong phage pressure. Of note, neither mucoid nor non-mucoid populations exhibited changes in ciprofloxacin susceptibility during first- or second-line therapy (Figure S9). Together, these findings highlight the complex interplay between bacterial genomic diversity, phenotypic adaptability, and therapeutic response in both acute and chronic *P. aeruginosa* infection contexts.

### Phage therapy alters lung virome through self-dosing dynamics

To assess therapeutic efficacy, we analyzed virome dynamics in patient sputum during treatment. Before intervention, the virome was dominated by *Litunavirus* phages (99% identity to PYO2), with lower levels of *Pakpunavirus* phages (∼30% similarity to P1A) and minor representation (2–3%) of non-*Pseudomonas* phages such as *Tequatrovirus* (Figure 4a, Tables S3, S7). Following phage therapy initiation, *Pakpunavirus* rapidly expanded from <25% to 98%, while *Litunavirus* declined to <2%, consistent with successful delivery and in situ amplification of P1A, as confirmed by near-complete genome identity. In contrast, PYO2 did not reach detectable levels in sputum. By day 9, P1A abundance decreased by ∼75%, coinciding with a marked drop in bacterial load (Figure 3a). At the same time, *Pbunavirus* phages expanded but were not associated with further bacterial clearance. Post-treatment, *Pakpunavirus* and *Pbunavirus* each comprised 37–45% of the virome, alongside the emergence of additional genera (Table S3), indicating sustained therapeutic phage presence and broader virome restructuring.

**Figure 4:**
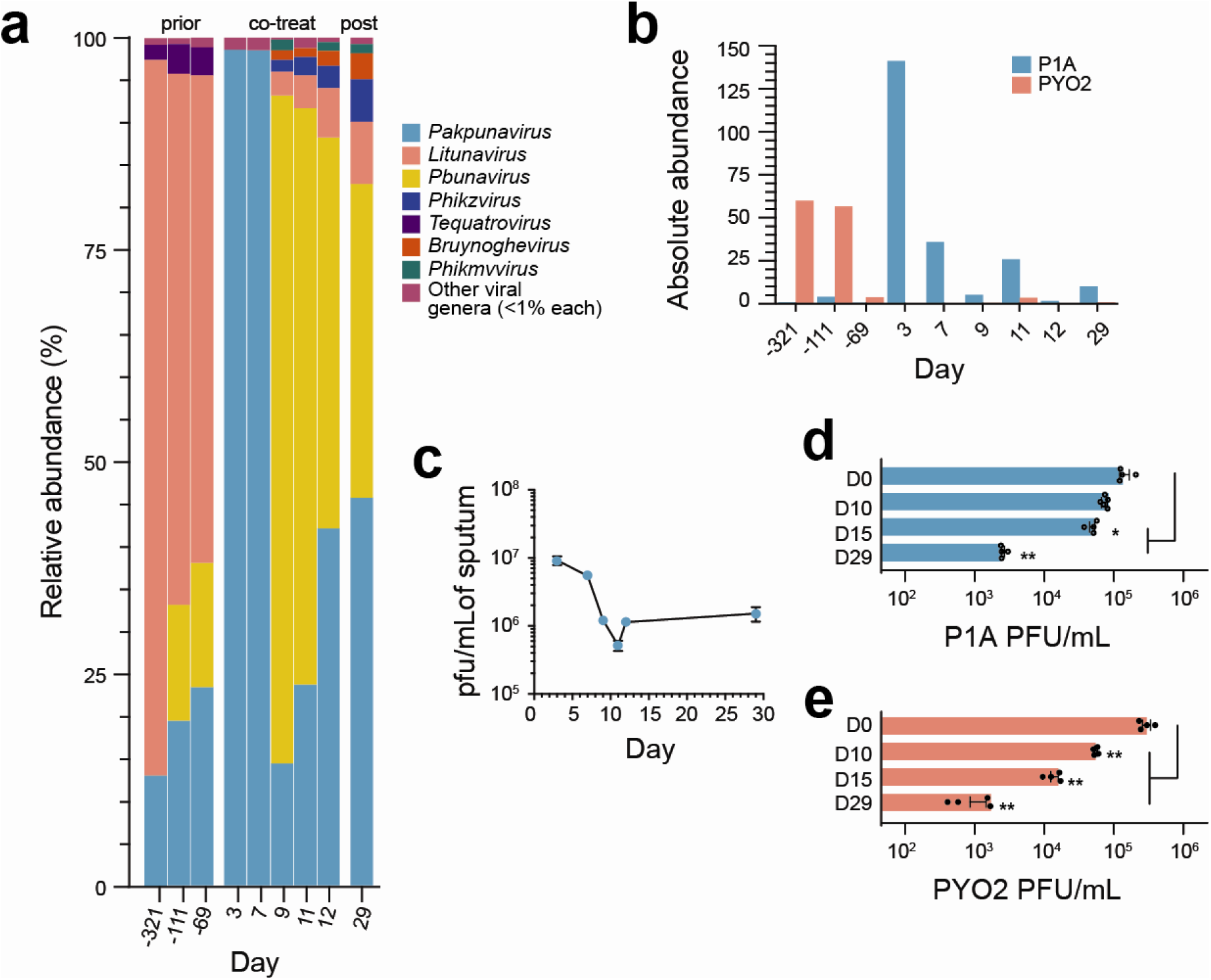
P1A demonstrated robust *in situ* replication following intravenous administration but was neutralized during its second week of treatment. (a) Stacked bar plots of relative abundance as fractions of total viral reads (%) before, during, and after co-therapy. Bars are colored by viral genera: *Pakpunavirus* (light blue), *Pbunavirus* (yellow), *Litunavirus* (pink), *Phikzvirus* (dark blue), *Tequatrovirus* (purple), *Bruynoghevirus* (orange), *Phikmvvirus* (green), and other taxa each with < 1% total reads (magenta). (b) Bar graph of absolute abundances of reads mapped to therapeutic phages P1A (blue) and PYO2 (pink). Absolute abundances were calculated by normalizing viral reads against diploid human genomes within each sample. (c) Estimated quantities of P1A plaque-forming units per mL (pfu/mL) in unenriched patient sputa. Phage titers were extrapolated from colony counts (Figure 3a) using the multiplicity of infection (MOI) of treatment phage P1A. The *in situ* MOI was calculated using the equation = (P1A absolute abundance)/(*Pseudomonas* absolute abundance), where each value was normalized for virions/bacteria per human cell. (d-e) Bar graphs showing *in vitro* phage neutralization of sera samples. Phages (approx. 10^5^ pfu/mL) (d) P1A and (e) PYO2 were co-cultured with 10% diluted archival sera and plaque formation was quantified after 24 h incubation. Asterisks indicate statistical significance **p < 0.01, *p < 0.05.

To evaluate in situ phage replication, we quantified the absolute abundance of therapeutic phages over time. PYO2-related phages peaked before the exacerbation at ∼60 virions per human cell (Figure 4b), but despite a spike in *Pseudomonas* abundance to 167 cells per human cell on day −69 (Figure 2d), endogenous phages did not expand, suggesting poor targeting of the exacerbation strain. During co-treatment, PYO2 levels remained low, indicating limited *in situ* amplification despite ongoing IV administration. In contrast, P1A was undetectable pre-treatment but rose to 141 virions per human cell within three days of therapy. From absolute abundance data, we estimated a local concentration of 9.2 × 10 PFU/mL and an MOI of 0.78 (Figure 4c vs. Figure 3a, Figure S10). Although P1A declined by day 7, concurrent reductions in bacterial load maintained a favorable MOI of 0.91 (Figure 4b–c). Given the IV dose alone could not account for the observed phage levels, this indicates substantial in situ replication. P1A abundance declined further in the second week, likely due to reduced host availability (Figure 3a), but maintained an MOI of 1.1 post-treatment, suggesting stable integration into the lung virome. These dynamics, combined with the observed expansion of *Pbunavirus* phages, some of which specifically targeted mucoid bacterial populations (Figure S9), suggest a shift toward a more balanced lung virome in which multiple distinct phage genera collectively contribute to controlling both nonmucoid and mucoid P. aeruginosa populations.

### Phage neutralization compromises cocktail efficacy

Phages can significantly influence the human immune system ^21^. Given the decline in P1A efficacy and the limited detection of PYO2 in sputum, we investigated whether humoral immunity contributed to therapeutic failure. Serum analysis revealed strong neutralizing responses against both phages. Neutralization of P1A became evident by day 15 (Figure 4d), consistent with expected antibody response timelines ^34^, and likely contributed to reduced lung abundance (Figure 4a–b) and diminished efficacy in the second week (Figure 3a). In contrast, robust neutralization of PYO2 appeared earlier, by day 10 (Figure 4e), suggesting pre-existing immunity, supported by prior detection of closely related *Litunavirus* phages with 99% genomic similarity (Figure 4b, Table S7). Immune activation was further supported by a mild increase in white blood cell count (Figure 1d). Over time, neutralization of PYO2 intensified, reducing its pulmonary availability by ∼3 logs (Figure 4e). These findings highlight the rapid and clinically significant impact of phage-specific antibody responses, indicating that while P1A was initially effective, PYO2 faced substantial immunological barriers.

## DISCUSSION

This study reframes both what is possible and what is necessary in clinical phage therapy for cystic fibrosis (CF). In an era where antimicrobial resistance continues to outpace therapeutic innovation, we demonstrate that phage and antibiotic co-therapy can drive rapid functional recovery, reduce bacterial burden, and reshape the lung microbiome and virome in a patient population largely excluded from phage trials: the elderly. Beyond its clinical success, this case reveals biological constraints that challenge prevailing assumptions about phage function in vivo and highlight critical priorities for future investigation.

The most immediate clinical implication is that phage therapy can be safely and effectively administered to late-stage, high-risk CF patients. The patient tolerated high-dose intravenous phage infusions without hypersensitivity or adverse events and experienced the most rapid and complete recovery from pulmonary exacerbation in over a decade. This outcome positions phage therapy not as a last-resort experimental option, but as a viable therapeutic intervention, particularly when conventional antibiotics fail. Importantly, this was not anecdotal: measurable improvements in forced vital capacity, spirometry, qCT-confirmed obstruction clearance, and PCA-derived health clustering all support the additive benefit of phage therapy when used alongside antibiotics.

What distinguishes this case is not just the outcome, but the mechanistic insight into how it was achieved. The multidrug-resistant non-mucoid *P. aeruginosa* population declined by three orders of magnitude within 24 hours, coinciding with a peak in in situ phage replication. This provides the strongest human evidence to date of phage self-dosing, a cornerstone of phage therapy’s theoretical advantage. Unlike antibiotics, phages amplified directly at the site of infection, driving a localized response as long as the infection persisted and remained susceptible. However, this effect was transient. By day 7, phage levels declined, and resistant bacterial subpopulations emerged. These resistance events were not biologically neutral. The resulting strains exhibited phenotypic alterations, including growth defects and hyperpigmentation, indicating that phage pressure induced evolutionary bottlenecks with potential consequences for pathogen fitness. Phage therapy should therefore be viewed not only as a bacteriolytic tool, but also as a strategy to steer pathogens toward less fit, less virulent states.

In contrast, the mucoid *P. aeruginosa* subpopulation, characteristic of chronic CF lung infections ^35^, remained largely unaffected. Its persistent resistance to both phages and antibiotics likely stemmed from biofilm-mediated protection rather than genetic resistance. This reinforces a critical but often clinically overlooked principle: bacterial susceptibility is shaped not only by genotype, but by phenotype and microenvironment. Without phage cocktails capable of penetrating biofilms, or formulations that disrupt the matrix barrier, a substantial portion of the CF lung microbiota will likely remain inaccessible to therapy ^36^. Inhaled mucolytics and biofilm-targeting phages should be considered essential components of future CF treatment regimens.

Equally consequential is the host’s role in shaping therapeutic outcomes. Within 10 to 15 days, both phages were neutralized by host immunity, coinciding with diminished efficacy. For PYO2, this likely reflected pre-existing immunity due to prior exposure to *Litunavirus*-like phages, while P1A elicited de novo antibody responses. These findings underscore a key limitation of systemic phage delivery. Although intravenous administration is clinically convenient, it may be unsustainable in patients with prior phage exposure or rapid immune activation. Baseline immune profiling should become standard for phage therapy in persistent infections, and alternative delivery strategies that avoid systemic immune priming, such as inhaled, mucosal, or encapsulated formulations, should be prioritized.

One of the more transformative findings in this study involves the virome. After therapeutic phage levels declined, non-treatment *Pbunavirus* phages expanded and persisted beyond the treatment window. Whether this shift contributed to *P. aeruginosa* suppression or reflected ecological rebalancing is unclear. Still, it reframes the virome as a potentially active participant in therapeutic response rather than a passive backdrop. Monitoring the bacterial microbiome alone is no longer sufficient; we must also consider how phage therapy reshapes the broader viral ecology of the lung, and whether these changes can be harnessed or intentionally guided. Virome shifts may serve as biomarkers of response ^37^, relapse ^38^, or resistance. Future studies should incorporate longitudinal, high-resolution virome surveillance as a central component of phage therapy assessment.

Together, these findings call for a redefinition of what constitutes success in phage therapy. Complete pathogen eradication is not the only meaningful endpoint; clinical benefit may also arise from reducing obstruction, shifting strain dominance, and leveraging selective pressure to induce fitness trade-offs. This challenges conventional reliance on culture conversion and underscores the need for more nuanced outcome measures. Future evaluations should incorporate functional lung metrics, track microbial evolution, and assess virome restructuring as indicators of therapeutic impact or durability.

Our findings also support several forward-looking hypotheses. First, that phage-induced resistance, though well documented in vitro ^39^, may provide clinical benefits when it imposes fitness costs such as impaired growth or virulence ^40^. Second, that systemic phage therapy is limited by host immunity unless personalized through immune profiling or reformulated to avoid neutralization. Third, that the lung virome is a dynamic and modifiable ecosystem that may influence treatment outcomes. Testing these hypotheses will require moving beyond conventional trial designs. Adaptive dosing ^41^, real-time genomic and virome monitoring, and integrated immune assessments should become core components. Randomized trials alone will be insufficient unless structured to capture the ecological and immunological complexity of phage therapy in practice.

Despite its depth, this study has several limitations. As a single-patient, compassionate-use case, it is not generalizable and cannot establish causality. The absence of a control arm limits the attribution of effects to phage therapy, despite strong temporal correlations. Sampling gaps, particularly pre-treatment, constrained baseline microbial and immune resolution. Immune profiling focuses on systemic responses, without assessing local immunity in the respiratory tract. Virome shifts, while striking, remain correlative. And although short-term recovery was substantial, long-term durability remains unknown. These gaps highlight the need for larger, prospective studies with standardized endpoints and extended follow-up.

In conclusion, this study represents an inflection point for the clinical application of phage therapy. It demonstrates not only that phage therapy can succeed in high-risk patients, but also clarifies when it works, why it fails, and what must be reimagined. To realize its full potential, phage therapy must evolve from anecdotal use to a precision-guided, ecologically informed, and immunologically integrated therapeutic approach. With the necessary tools and models now in place, what remains is the strategic vision to bring them into clinical practice.

## MATERIALS & METHODS

### Ethics approval and consent

Written informed consent was obtained prior to any study-specific procedures, with re-consent obtained as necessary. The study protocol and subsequent amendments were approved by the FDA (IND #26965), with local approval granted by the Institutional Review Board of UC San Diego (IRB #802083). Treatments were administered inpatient via IV twice daily through a peripherally inserted central catheter (PICC) line.

Following discharge, the patient continued self-administration of phages through the PICC line, demonstrating high adherence to the prescribed phage therapy.

### Patient health metrics and pulmonary function tests

The patient was seen at UC San Diego Medical Hospital for all pulmonary function and hematology tests. Clinical microbiology, hematology, and x-rays were performed during normal care and clinical follow-ups of the patient. At each visit the patient’s height, weight, body mass index, urine, and spirometry were collected. Spirometry measurements were conducted according to American Thoracic Society guidelines and normalized using the Hankinson equations ^42^. Forced expiratory volume in 1 second (FEV_1_), forced vital capacity (FVC), and maximum mid-expiratory flow (MMEF) values were collected. FEV_1_ and FVC tests values were then used to calculate the percentage of predicted (pp) to obtain ppFEV_1_ and ppFVC as previously described ^43^. Quantitative computed tomography was likewise performed at UCSD Medical Hospital, and an automated CT algorithm was used for CT quantification of % emphysema, % air trapping, inspiratory volume, and expiratory volume as previously described ^44^. Urine samples were used to measure creatinine levels.

### Bacterial strains and isolate collection

*Pseudomonas aeruginosa* laboratory strains PAK (CP020659.1) and PAO1 (NC_002516.2) were used to propagate phages P1A and PYO2, respectively. Laboratory strains were cultured overnight in Luria broth (LB) at 37°C with shaking. Clinical *P. aeruginosa* isolates were obtained from patient sputum samples using selective cetrimide agar and were subsequently streak-purified to isolate single colony morphologies. Patient bacterial isolates were then grown in LB at 37°C with shaking for *in vitro* sensitivity testing and DNA isolation. Isolates are named for their isolation date, relative to the first day of phage treatment (day 0, D0). Isolates were stored in 25% (v/v) glycerol at −80°C.

### Phage strains and clinical production

Phages P1A and PYO2 (MF490236.1) were confirmed not to contain known toxins, harmful and antibiotic resistance genes, as well as lysogeny-related mechanisms. Each phage’s enrichment was performed by inoculating bacterial cultures with an MOI of 0.01 to yield a 3L lysate. The amplified phages were sterilized via double centrifugation and filtration at cutoffs of 0.8, 0.45, and 0.22 µm. The 3L lysate was concentrated using a 100 kDa Vivaflow cross-flow filtration cassette (Sartorius), further removing endotoxins, exotoxins, and other potential contaminants, and then diafiltered with sterile cold phosphate buffer saline (PBS) before final concentration to >10^11^ pfu/mL. Intact phage particles were further separated with cesium chloride (CsCl) density-gradient ultracentrifugation at 100,000 × g for 12 hours, followed by 3 rounds of dialysis in fresh cold PBS to remove CsCl. Final LPS scrubbing was performed with Pierce™ High Capacity Endotoxin Removal Spin Columns.

### Phage quantification

Phage concentrations were monitored throughout the purification process and for *in vitro* experiments using the spot titering method ^45^. For each sample, tenfold serial dilutions of phage stock were prepared in a microplate and spotted onto bacterial-seeded agar plates.

### Phage strain characterization

Measurements for phages P1A and PYO2 were conducted on respective isolation hosts PAK and PAO1. The rate of adsorption (binding of individual phage particles) was measured by calculating the decrease in free- phages in culture with host cells ^46^. Approximately, phages were diluted into warm growth medium (1 × 10^7^ PFU) and used to inoculate host cells (4.5 × 10^8^ CFU) at 37°C with shaking (120 rpm). At 1 min intervals 100µL samples were aliquoted, treated with chloroform, and centrifuged at 12,000×g for 3 min. Each timepoint was titered using the spot method. The one-step phage growth curve was measured as described by Kropinski ^47^. Phages were diluted (5 × 10^7^ PFU) in warm growth broth and used to inoculate host cells (4.95 × 10^8^ CFU) at a MOI 0.1 and incubated at 37°C with shaking (120 rpm). Duplicate samples were taken at 2 min intervals, treated with and without chloroform, and centrifuged at 12,000×g for 3 min. Phages from each timepoint were titrated using the spot method. Both phage adsorption assays and one-step growth curves were each performed with three isolated CFUs.

### Phage stock characterization

Phage master stocks were determined by spot titration:

- P1A: 5.88 × 10^11^ pfu/mL containing 453 endotoxin units (EU)/mL
- PYO2: 5.81 × 10^11^ pfu/mL containing 8920 EU/mL

Phage strain identity was confirmed by PCR with strain specific primers. The two phages were mixed to create individual therapeutic doses at 2×10 pfu (1×10 pfu per strain) in 1 mL of certified pyrogens free Dulbecco’s phosphate buffered saline (dPBS). The solution was 0.22 µm filtered, sterile fill-finished into single dose sealed glass vials, stored protected from UV, and transported at 4°C.

### Sterility and endotoxin testing

FOCUS Laboratories (Allentown, PA, USA) confirmed that each phage production batch met sterility standards (USP 71) and determined (USP 85) that each phage dose contained <1 EU/mL/kg of patient body weight (Figure S3).

### Phage cytotoxicity testing

The cytotoxicity of each phage was assessed using the CellTiter-Glo® 2.0 Assay (Promega) as described ^45^. A549 (ATCC CCL-185) lung epithelial cells were cultured in Dulbecco Modified Eagle Medium (DMEM) with 10% fetal bovine serum (Gibco) at 37°C in 5% CO_2_. Cells were seeded in a microwell phage at ∼20,000/well and incubated for 24 h. Purified phage stocks were diluted to 10 pfu/mL and added to achieve a 1:1000 ratio of cells to phages. SDS (1%) was used as a positive control and PBS as a negative control. After a further 24 h incubation, CellTiter-Glo was added, and the plate was mixed for 2 min. Following a 10 min incubation at 22°C, luminescence was measured using a microplate reader (BMG Labtech).

### Antibiotic susceptibility testing

Sputa sample colonies were grown and tested for susceptibility by the UCSD Clinical Microbiology using VITEK® 2 (BioMérieux) for most antibiotics and disk diffusion for ceftalozone-tazobactam. MIC cutoffs were based on CLSI breakpoints (2022). Further ciprofloxacin sensitivity testing of isolated non-mucoid and mucoid isolates from patient sputa was tested in liquid culture at a range of 16 µg/mL to 0.03 µg/mL. MIC was determined by the lowest concentration with no bacterial growth, indicated by an OD600 <0.3.

### Sputum collection and DNA extraction

Spontaneously expectorated sputum was collected in a sterile container and briefly stored at 4°C until DNA extraction. The freeze-boil method described by Silveira et al. ^48^ was used for metagenomic DNA extraction. In short, homogenized sputum was stored in a temperature-resistant cryovial and subjected to five 5 min freeze-boil cycles (-80°C and 100°C, respectively). After liquefaction, the sample was centrifuged to remove debris, and the supernatant was processed using the NucleoSpin Microbial DNA Isolation Kit (Machery-Nagel, Düren, Germany) according to the manufacturer’s protocol.

### Sequencing, metagenomic taxonomic classification, and metagenomic abundances

Metagenomic DNA libraries were prepared by SeqCenter (Pittsburgh, PA, USA) using the Nextera DNA Flex sample preparation kit (Illumina) for Illumina NextSeq 550 paired-end sequencing. For taxonomic classification, paired-end reads were processed with *fastp* (v0.19.4) ^49^ and then classified using *Kraken2* (v2.1.3) ^50^ with default settings. Taxonomic results were visualized using *Pavian* (v1.0) ^51^ to determine classified and unclassified reads for each taxon. Relative abundance was calculated as the ratio of reads per taxon to the total classified reads. Absolute abundance was determined following Pust et al. ^25^, scaling the diploid human genome length to millions, multiplying by the normalized bacterial read count (per million reference base pairs), and dividing by the human read count.

### Data standardization and analysis

Variables including FEV_1_, FVC, FEV_1_/FVC, ppFVC, ppFEV_1_, MMEF, creatinine, WBC, and platelets were standardized to have a zero mean and unit variance in preparation for principal component analysis (PCA). PCA was conducted using the *prcomp* function with centering and scaling to reduce the dataset to principal components that capture the most important patterns and variance. The first two principal components were visualized using the *ggplot2* package (v. 3.4.3). Hierarchical clustering analysis (HCA) was performed using the *hclust* function, based on a distance matrix derived from PCA scores. Ward’s method (ward.D2) was applied to minimize within-cluster variance by squaring dissimilarities before updating clusters ^52^. All PCA and dendrogram functions were sourced from the R stats package (version 3.6.2). Statistical analyses followed SAS standards.

### Quantification of total microbial load

Sputum samples were archived with 50% glycerol and stored at −80°C until colony quantification. Archival sputum glycerol stocks were scraped to obtain 50μL samples which were further serially diluted in sterile dPBS. After dilution, 650μL of each dilution series was spread onto Luria broth (LB) agar plates and incubated at 37°C for 24 h. Total CFU’s were counted and were categorized into non-mucoid and mucoid phenotypes. Plating was repeated in triplicate for each sputum sample date (n=3).

### Pangenome phylogenies

Microbial DNA were extracted from cultures of individually purified patient bacterial isolates using the NucleoSpin Microbial DNA Isolation Kit according to manufacturer’s protocol. Sequencing was prepared likewise to metagenomes by SeqCenter (Pittsburgh, PA, USA). For bacterial isolate assembly, the paired-end reads were processed with *fastp* (version 0.19.4) ^49^ with default settings. High-quality reads were *de novo* assembled into contigs with SPAdes (v4.0.0) ^53^ using default parameters. The resulting assemblies were annotated using PAO1 (NC_002516.2) as a reference genome with *Prokka* (v. 1.14.5) ^54^ for gene prediction and functional annotation. We separated the sequences based on their mucoid and non-mucoid phenotypes to perform subpopulation core genome alignments using *Roary* (v. 3.11.2) ^55^. Two sets of trees were created: a core genome phylogeny was generated using the core genome alignments and an accessory genome phylogeny was generated using a binary gene presence absence comparison. For each alignment, a maximum-likelihood phylogeny tree was created using FastTree ^56^ with a generalized time-reversible (GTR) model with 1000 iterations. The resulting trees were visualized using iTOL (Interactive Tree of Life) (v. 6.0.0) ^57^.

### Phage susceptibility testing

For susceptibility testing, 4 µL of phage concentrate (10 and 10^7^pfu/mL) was spotted onto bacterial-seeded agar plates. After allowing spots to dry for approximately 15 min, the plates were incubated at 37°C for 24 h. The presence or absence of plaques was recorded for each bacterial isolate.

### Time-kill kinetics

Non-mucoid isolates from clade 4 lineage were grown overnight in LB at 37°C. Replicates (n=3) were initiated from independent CFUs, pelleted, and resuspended in sterile Milli-Q water. In a 96-well microplate 2× LB and 2 × 10 CFU were added. The plate was incubated at 37°C with orbital shaking in a microplate reader (BMG Labtech) for 24 h, with absorbance taken every 6 min. Bacterial densities were quantified using the spot method. Area under the curve was calculated by fitting OD_600_ data using the *Growthcurver* R package ^58^.

### Phage strain mapping to metagenomes

In order to specifically map therapeutic phages P1A and PYO2 to metagenome reads, *bbmap* ^59^ was used to align complete phage genomes to the plus and minus reads after quality control by *fastp*. A minimum alignment identity cut-off of 0.95 was used (minid=0.95).

### Serum preparation and neutralization testing

Phages P1A and PYO2 were diluted to 1 × 10 pfu/mL in sterile dPBS. Collected serum was diluted in sterile dPBS to achieve a final concentration of 10%. Phages were then added to the serum to reach a final concentration of 1 × 10 pfu/mL. The phage-serum mixture was incubated at room temperature for 2 and 24 hours. After each incubation period, phage concentrations were determined by serial dilution and plating onto their respective isolation host strains using the spot plaque titer assay. Plates were incubated overnight at 37°C, and plaques were counted to quantify phage recovery.

### Data and code availability

All data produced in the present study are available upon reasonable request to the authors. User scripts will be shared upon request.

## Acknowledgments

We thank G. Burkeen for assistance with sample handling, J. Mielke for administrative support, R. Cook for bioinformatics guidance, and F. Rohwer for valuable feedback on the study. This work was supported by an ARCS Foundation Scholarship (TL), the Cystic Fibrosis Foundation (KCJ), and the Conrad Prebys Foundation (DRR).

## Author Contributions

Conceptualization, TL, DC, and DRR; methodology, TL, LK, KC-J, DJC, and DRR; software/code, TL and LK; validation, TL and DRR; investigation, TL, LK, KC-J, JAM, and DJC; formal analysis, TL, LK, KC-J, DJC, and DRR; resources, DP, DJC, and DRR; data curation, TL and LK; original draft, TL, LK, DJC, and DRR; review & editing, TL, DC, and DRR; visualization, TL, LK, and DRR; supervision, DJC and DRR; funding, DJC and DRR.

## List of Supplementary Data

Figures S1, S2, and S3 supporting Figure 1.

Tables S1 and S2 supporting Figure 1.

Tables S3, S4, S5, and S6 supporting Figure 2.

Figures S4, S5, S6, S7, S9 supporting Figure 3.

Table S7 supporting Figure 4.

Figure S10 supporting Figure 4.

