## Supplemental Figures S1-S10 for "Ecological and Immune Pressures Shape Outcomes of Precision Phage Therapy in Advanced Cystic Fibrosis Lung Disease"

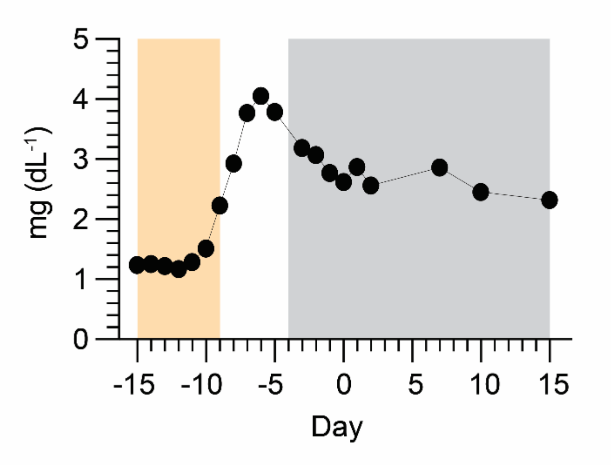


**Figure S1: Colistin administration dangerously increased the patient's baseline creatinine levels by 4-fold.** Line graph of patient urine creatinine concentration (mg·dL$^{-1}$) which increased in response to first-line colistin treatment (days -15 to -9, yellow box), climbing to a peak of 3.79 mg·dL$^{-1}$ on day -5. Urine creatinine exceeding 1.3 mg·dL$^{-1}$ indicates improper kidney filtration and function in an adult male (Shahbaz and Gupta 2023). Creatinine levels began receding on day -4, five days after the antibiotic was stopped. Administration of second-line treatment, staggered starting with ciprofloxacin on day -4 and phage addition on day 0, did not lead to a resurgence in creatinine levels.

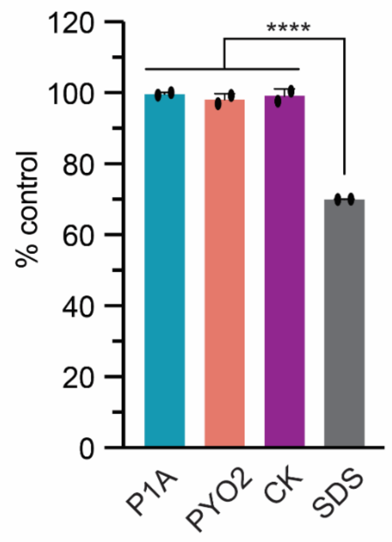


**Figure S2: Purified phage preparations are non-toxic to *in vitro* lung epithelial cell culture.** CellTiter-Glo 2.0, an assay commonly used for testing the viability of cells in response to therapeutic treatments, was used to confirm non-toxicity of phage preparations against human lung epithelial cells (A549). Phages were individually purified (see Methods) and individual phage strains *Pakpunavirus* P1A (blue) and *Litunavirus* PYO2 (pink) as well as the formulated phage cocktail (purple) were tested. Cell responses were normalized to a PBS (no phage) control and the detergent sodium dodecyl sulfate (SDS) was used as a positive cell lysis control.

FOCUS LABORATORIES

### Certificate of Analysis No. 22-00057

| | | | |
|---|---|---|---|
| Customer: | University of California San Diego | Contact: | |
| Address: | 9500 Gilman Drive, La Jolla<br>California, 92093, | Contact Email: | |
| Date Authorized: | | Total Number of Pages: | 1 |

#### Test Results

| | | | |
|---|---|---|---|
| Sample No. | 22-00057-01 | Sample Received Date: | |
| Product Name: | P.aeruginosa Phage Cocktail P1A \| PYO2 | Sample Description: | Concentration: 2 x 10^8 PFU/mL |
| Lot Number: | PsA76.2 | | |
| Comments: | Volume tested for Scan RDI: 8mL | | |

| Test | Method | Results | Specification | Pass/Fail | Completed | Lab |
|---|---|---|---|---|---|---|
| Endotoxin (EU/mL) | USP <85> | <1 EU/mL | <370 EU/mL | PASS | | FOCUSFL |
| Sterility by Scan RDI | FOCUS SOP 1401.01 | 0 microbes | <1 microbes | PASS | | FOCUSFL |

Reference Information:

Laboratories:

FOCUSFL : Focus Laboratories Florida, 2660 Alternate 19 N, Suite B, Palm Harbor, FL 34683

Reviewed / Approved by:

QA Coordinator

Signed Date:

FOCUS Laboratories Pennsylvania is ISO/IEC 17025:2017 accredited by PJLA in the field of testing, Accreditation #77499

†indicated specific test is outside the scope of ISO accreditation, however FOCUS is an FDA registered cGMP laboratory.
This document is issued under the company's terms and conditions which were agreed upon during sample submission. FOCUS Laboratories' liability regarding this analysis is limited to the cost of the tests associated with this Test Report. The results are the property of the client listed above and FOCUS Laboratories. These results only relate to items tested above and apply to the samples as received. This report shall not be reproduced except in full without approval of FOCUS Laboratories.

Page 1 of 1

**Figure S3: Highly purified phages produced in-house at SDSU meet safety criteria for intravenous administration.** Certificate of analysis for *P. aeruginosa* Phage Cocktail P1A | PYO2. FOCUS Laboratories, an FDA registered contract testing laboratory performed testing as per Food and Drug Administration (FDA) regulations. US Pharmacopeia (USP) 85 testing determined that the cocktail's endotoxin level (endotoxin units (EU)) was far below the minimum requirements as adjusted for the patient's weight. Because phages are a filterable (sterile filter of 0.2µm) drug product, ScanRDI was used with viability dye to rapidly identify the presence of microbes using same day testing. No microbes were detected in the phage preparation.

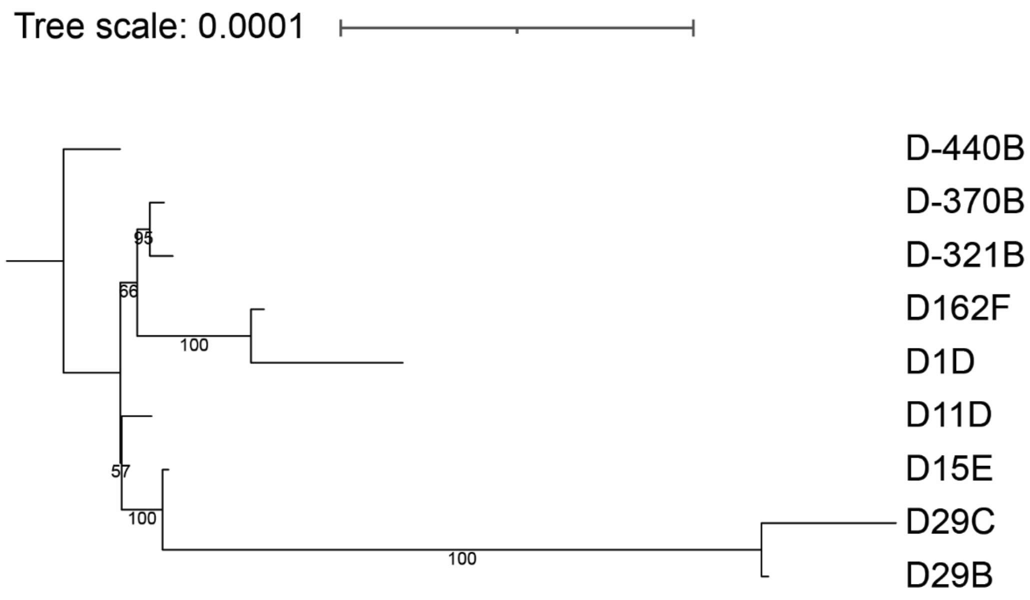


**Figure S4: Core genome phylogeny of mucoid *P. aeruginosa* exhibits low divergence within a 1.5 year-long span.** Maximum likelihood phylogenetic tree of mucoid *P. aeruginosa* subpopulation generated from core genome alignment using *Roary*. The tree was made with FastTree using a generalized time-reversible (GTR) model with 1000 iterations. The scale bar represents 0.0001 nucleotide substitutions per site and bootstrap values are shown at each node. The tree is rooted to the oldest mucoid isolate (D-440B) showing the relationship between 10 mucoid isolates sampled over time. The maximum distance between isolates was 0.00026 nucleotide substitutions per site between nodes at D-440B and D29C. Isolates exhibited low divergence and appeared to largely cluster into two clades with the first clade containing most of the older isolates (D-440B, D-370B, D-321B, D162F, D1D) and a second clade with the majority of post-treatment isolates (D11D, D15E, D29C, D29B). This would be consistent with a core genome requiring long periods of time (>1 year) to evolve during the patient's chronic infection.

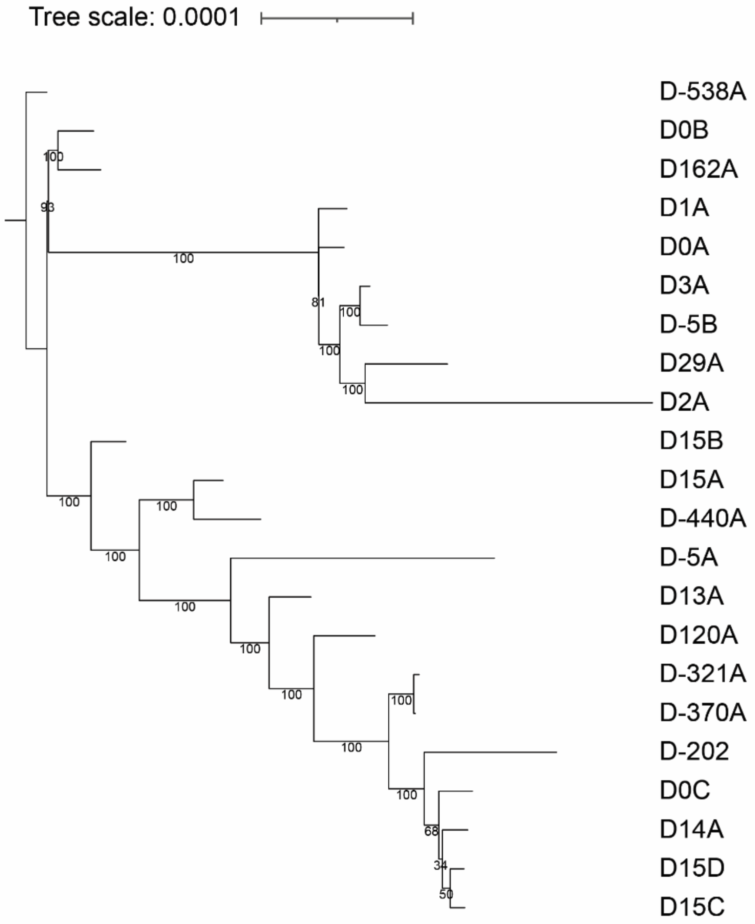


**Figure S5: Core genome phylogeny of non-mucoid *P. aeruginosa* shows limited divergence after phage therapy.** Maximum likelihood phylogenetic tree of clinical non-mucoid *P. aeruginosa* subpopulation made from *Roary* generated core genome alignment. The tree was made with FastTree using a generalized time-reversible (GTR) model with 1000 iterations. The scale bar represents 0.0001 nucleotide substitutions per site and bootstrap values are shown at each node. The tree is rooted to the oldest non-mucoid isolate (D-538A) showing the relationship between 22 non-mucoid isolates sampled over time. The maximum distance between isolates was 0.0004 nucleotide substitutions per site between nodes at D-538A and D2A. Isolates were more divergent than the mucoid tree (Fig. S4), likely due to the greater number of isolates used for tree generation but were likewise tightly clustered with relatively short branch lengths. Non-mucoid isolates diverged into 3 main clusters with a mixture of time points in each group consisting of one small group (D-538A, D0B, D162A), a medium-sized group (D1A, D0A, D3A, D-5B, D29A, D2A), and a large group of nested isolates (D15B, D16A, D-440A, D-5A, D13A, D120A, D-321A, D-370A, D-202, D0C, D14A, D15D, D15C). We surmise that this core phylogeny appears highly nested due to low divergence among the isolates, making separation of strains using only core genes difficult. The greatest genetic distance between isolates retrieved >500 days apart would again be consistent with the length of time required for diverging between isolates of the chronic infection.

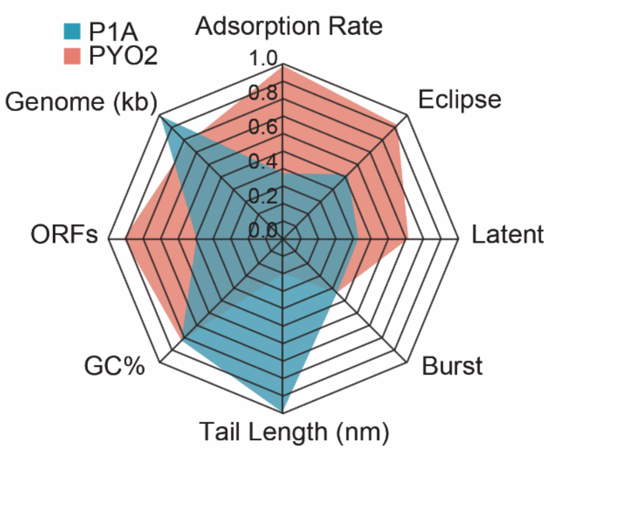


**Figure S6: Lytic phages P1A and PYO2 exhibit high replication activity within 30 minutes of infecting their respective isolation hosts.** Comparison of genomic and phenotypic properties of P1A (blue) and PYO2 (pink) on their isolation hosts. Phage infection involves virion acquisition of host cells (adsorption), creation of new virions (eclipse period), the infection itself (latent period), and the subsequent release of progeny virions from infected cells (burst). PAK_P1 adsorbs to the host cell surface over twice as fast as PYO2 ($3.79\times10^{-10}$ vs $1.61\times10^{-10}$ mL min$^{-1}$). Although, PAK_P1 has a shorter 15 min latent period compared to 25 min of PYO2, both strains produce and release about 100 progeny virions per infected cell. These phage parameters are comparable to the phenotypic features measured in other genera of virulent anti-pseudomonal phages (Ceyssens, Glonti et al. 2011, Forti, Roach et al. 2018). Genomic information was obtained by short-reading sequencing P1A and for PYO2 from RefSeq under accession number MF490236.1.

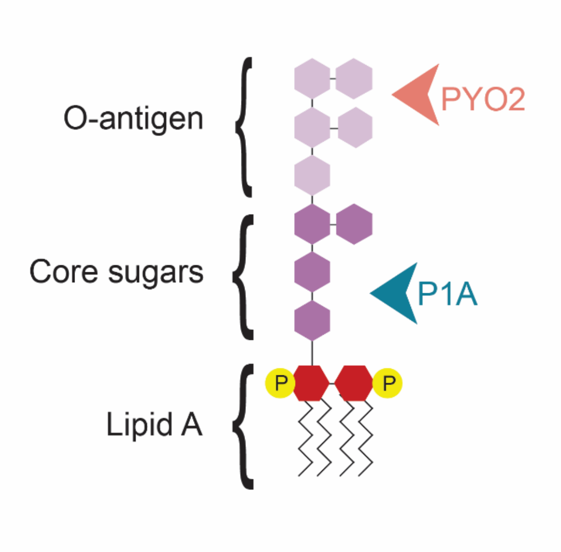


**Figure S7: Phages P1A and PYO2 bind to different *P. aeruginosa* lipopolysaccharide regions.** Schematic of lipopolysaccharide (LPS) binding patterns of P1A and PYO2. P1A and PYO2 bind to the inner core and O-antigen of LPS, respectively. Phage receptor binding patterns were determined via sequencing and single nucleotide polymorphism (SNP) analyses of phage resistance mutants (Forti, Roach et al. 2018, Schumann, Sue et al. 2022).

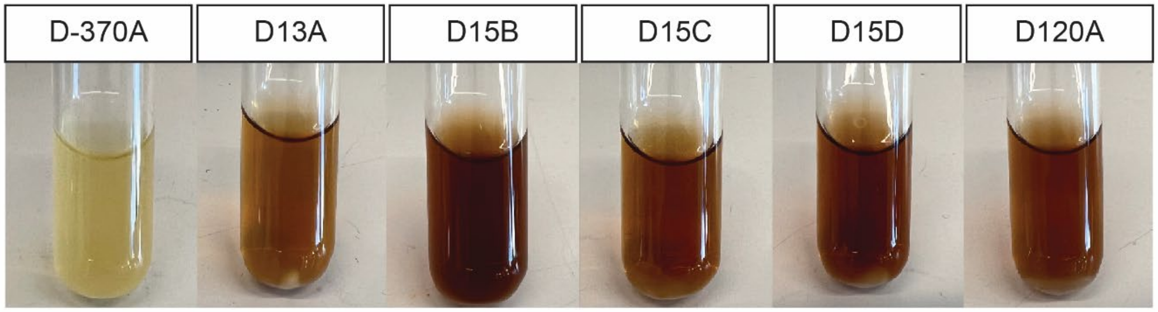


**Figure S8: Resistance to P1A causes bacterial expression of reddish-brown hyperpigmentation.** Patient non-mucoid isolates with resistance to phage P1A were inoculated from purified single colony-forming units. Each 5mL culture was incubated in LB media for 48 hours at 37°C with shaking at 120RPM. Test tubes are arranged chronologically and labeled corresponding to Fig. 3d nomenclature.

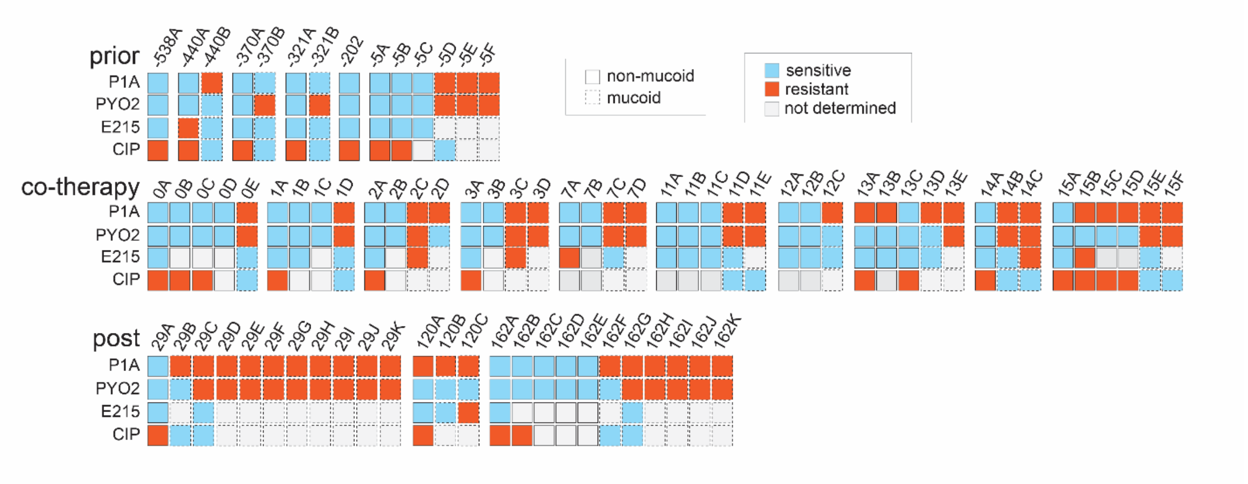


**Figure S9: Susceptibility to ciprofloxacin remained unchanged in both mucoid and non-mucoid populations throughout treatments.** Chronologically organized heatmap of clinical isolate susceptibilities to phages P1A, PYO2, E215, and ciprofloxacin. Phage E215 (RefSeq accession number: NC_042080.1) is a representative phage strain of the *Pbunavirus* genus, for viral characteristics refer to Forti et al. (Forti, Roach et al. 2018). Each column represents a single isolate, labeled by sputa sample date. Non-mucoid isolates are outlined with solid black and mucoid isolates are outlined with dashed black lines. For phages, the ability to plaque on a solid bacterial lawn at $10^8$ pfu/mL was considered sensitive (blue) and a lack of any clearance insensitive (orange). For ciprofloxacin, a minimum inhibitory concentration (MIC) cutoff between 0-2µg/mL was considered sensitive (blue) while MIC >2µg/mL was resistant (orange).

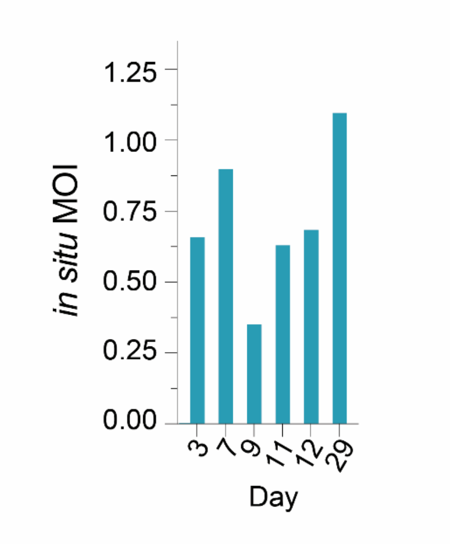


**Figure S10: Phage P1A concentration waxed and waned according to bacterial concentrations during co-treatment.** Bar graph of *in situ* multiplicity of infection (MOI) of treatment phage P1A after IV phage administration. MOI was calculated using the equation: *in situ* MOI = (P1A absolute abundance)/(*Pseudomonas* absolute abundance), where each value was normalized for reads per human cell.

### Figure References

Ceyssens, P. J., T. Glonti, N. M. Kropinski, R. Lavigne, N. Chanishvili, L. Kulakov, N. Lashkhi, M. Tediashvili and M. Merabishvili (2011). "Phenotypic and genotypic variations within a single bacteriophage species." Virol J **8**: 134.

Forti, F., D. R. Roach, M. Cafora, M. E. Pasini, D. S. Horner, E. V. Fiscarelli, M. Rossitto, L. Cariani, F. Briani, L. Debarbieux and D. Ghisotti (2018). "Design of a broad-range bacteriophage cocktail that reduces *Pseudomonas aeruginosa* biofilms and treats acute infections in two animal models." Antimicrob Agents Chemother **62**(6).

Schumann, A. R., A. D. Sue and D. R. Roach (2022). "Hypoxia Increases the Tempo of Phage Resistance and Mutational Bottlenecking of Pseudomonas aeruginosa." Frontiers in Microbiology **13**.

Shahbaz, H. and M. Gupta (2023). Creatinine clearance. StatPearls [Internet], StatPearls Publishing.
