## Supplemental Tables S1-S7 for "Ecological and Immune Pressures Shape Outcomes of Precision Phage Therapy in Advanced Cystic Fibrosis Lung Disease"

**Table S1: Patient health history and medication list.**

| Medical conditions | Antibiotic | Reaction | Medication list |
|---|---|---|---|
| Chronic sinusitis | Ciprofloxacin/levofloxacin | Severe tendonitis | Allopurinol |
| Exocrine pancreatic insufficiency | Meropenem | Non-anaphylaxis/diffuse rash | Inhaled aztreonam |
| Gout | Piperacillin/tazobactam | Non-anaphylaxis/diffuse rash | Famotidine |
| Osteopenia | Aminoglycosides | Hearing loss, vertigo, chronic kidney disease | Inhaled corticosteroids/long-acting beta2-agonists |
| Renal insufficiency with eGFR 40mL/kg/hr | Colistin | Chronic kidney disease | Atrovent |
| H/O Pneumothorax | | | Metoprolol |
| H/O hemoptysis | | | Pancreatic enzyme replacement therapy |
| Paroxysmal atrial fibrillation | | | Testosterone |
| Hearing loss with hearing aids | | | Elexacaftor/tezacaftor/ivacaftor |

**Table S2: Antibiotic minimum inhibitory concentration (µg/mL) susceptibility of *P. aeruginosa* sputa isolates.**

| Sample Date | Morphotype[2] | Antibiotic[1]: Amikacin[3] | | Aztreonam[3] | | Cefepime[4] | | Ceftazidime[4] | | Ceftazidime/Avibactam | | Ceftolozane/Tazobactam | | Ciprofloxacin[4] | | Colistin[3] | | Gentamicin[3] | | Levofloxacin[4] | | Meropenem[4] | | Piperacillin/Tazobactam[5] | | Tobramycin[3] | |
|---|---|---|---|---|---|---|---|---|---|---|---|---|---|---|---|---|---|---|---|---|---|---|---|---|---|---|---|
| D-538 | Mucoid | 4 | S | <=2 | S | 8 | S | 2 | S | 8/4 | S | <=2/4 | S | ND | ND | <=2 | S | 2 | S | ND | ND | <=0.5 | S | <=4/4 | S | ND | ND |
| | Nonmucoid | 16 | S | >16 | R | >16 | R | >16 | R | 16/4 | R | >8/4 | R | >2 | R | <=2 | S | >8 | R | ND | ND | >8 | R | >64/4 | R | <=2 | S |
| D-440 | Mucoid | 8 | S | <=2 | S | 8 | S | 2 | S | <=2/4 | S | <=2/4 | S | >2 | S | <=2 | S | 2 | S | >4 | R | <=0.5 | S | 8/4 | S | ND | ND |
| | Mucoid | 4 | S | <=2 | S | 4 | S | 2 | S | <=2/4 | S | <=2/4 | S | ND | ND | <=2 | S | ND | ND | 4 | I | <=0.5 | S | <=4/4 | S | ND | ND |
| D-370 | Mucoid | 8 | S | <=2 | S | 8 | S | 4 | S | <=2/4 | S | <=2/4 | S | >2 | S | <=2 | S | 4 | S | ND | ND | <=0.5 | S | <=4/4 | S | ND | ND |
| | Nonmucoid | >32 | R | >32 | R | >32 | R | >32 | R | 8/4 | S | >8/4 | R | ND | ND | <=2 | S | >8 | R | ND | ND | 4 | S | >64/4 | R | ND | ND |
| D-321 | Mucoid | <=4 | S | <=4 | S | 8 | S | <=2 | S | ND | ND | ND | ND | >2 | R | ND | ND | 2 | S | ND | ND | <=1 | S | <=8 | S | <=1 | S |
| | Nonmucoid | >32 | R | >16 | R | >16 | R | >16 | R | >16/4 | R | >8/4 | R | >2 | R | <=2 | S | >8 | R | ND | ND | 2 | S | >64/4 | R | 8 | I |
| D-69 | Mucoid | <=4 | S | >16 | R | 16 | I | 4 | S | ND | ND | ND | ND | 2 | I | ND | ND | <=1 | S | ND | ND | <=1 | S | <=8 | S | <=1 | S |
| | Nonmucoid | >32 | R | >16 | R | >16 | R | >16 | R | 16/4 | R | >8/4 | R | >2 | R | <=2 | S | >8 | R | ND | ND | 8 | R | >64/4 | R | 8 | I |
| D-13 | Mucoid | >32 | R | >16 | R | >16 | R | >16 | R | ND | ND | ND | ND | 2 | I | ND | ND | >8 | R | ND | ND | 8 | I | >64 | R | 8 | I |
| | Nonmucoid | 32 | I | >16 | R | >16 | R | >16 | R | 16/4 | R | >8/4 | R | >2 | R | <=2 | S | >8 | R | ND | ND | >8 | R | >64/4 | R | 4 | S |
| D1 | Mucoid | 8 | S | <=4 | S | 8 | S | 8 | S | ND | ND | ND | ND | >2 | R | ND | ND | 4 | S | ND | ND | 4 | S | <=8 | S | <=1 | S |
| | Nonmucoid | 32 | I | >16 | R | >16 | R | >16 | R | >16/4 | R | >8/4 | R | >2 | R | <=2 | S | >8 | R | ND | ND | >8 | R | >64/4 | R | 4 | S |
| D78 | Mucoid | <=4 | S | <=4 | S | 8 | S | <=2 | S | ND | ND | ND | ND | >2 | R | ND | ND | 2 | S | ND | ND | <=1 | S | <=8 | S | <=1 | S |
| D120 | Mucoid | 8 | S | 8 | S | 16 | I | >16 | R | ND | ND | ND | ND | >2 | R | ND | ND | 2 | S | ND | ND | 8 | I | <=8 | S | <=1 | S |
| | Nonmucoid | 16 | S | 8 | S | >16 | R | >16 | R | ND | ND | ND | ND | >2 | R | ND | ND | 4 | S | ND | ND | >8 | R | 16 | S | <=1 | S |
| D176 | Mucoid | 8 | S | <=4 | S | 8 | S | 4 | S | ND | ND | ND | ND | >2 | R | ND | ND | 2 | S | <=1 | S | <=1 | S | 32 | S | <=1 | S |
| | Nonmucoid | 16 | R | >32 | R | >32 | R | >32 | R | >16/4 | R | >8/4 | R | ND | ND | <=2 | S | >8 | R | 8 | I | 8 | I | >64/4 | R | ND | ND |
| D281 | Mucoid | 4 | S | <=2 | S | 2 | S | <=0.5 | S | <=2/4 | S | <=2/4 | S | ND | ND | <=2 | S | 1 | S | 1 | S | <=0.5 | S | <=4/4 | S | ND | ND |
| D358 | Mucoid | <=2 | S | <=2 | S | 2 | S | <=0.5 | S | <=2/4 | S | <=2/4 | S | ND | ND | <=2 | S | 2 | S | ND | ND | 1 | S | <=4/4 | S | ND | ND |
| | Nonmucoid | 32 | I | >16 | R | >16 | R | >16 | R | >16/4 | R | >8/4 | R | >2 | R | <=2 | S | >8 | R | ND | ND | >8 | R | >64/4 | R | 4 | S |

[1] Susceptibility interpretations are based on Clinical and Laboratory Standards Institute (CLSI) breakpoints.

[2] Isolate morphology determined on blood agar plate culture.

[3] Low risk of *C. difficile* infection.

[4] High risk of *C. difficile* infection.

[5] Moderate risk of *C. difficile* infection.

Abbreviations: ND, not determined.

**Table S3: Sputum metagenome reads classified by Kraken2.**

| | Sample Date | | | | | | | | |
|---|---|---|---|---|---|---|---|---|---|
| **Taxonomic classification** | **D-321** | **D-111** | **D-69** | **D3** | **D7** | **D9** | **D11** | **D12** | **D29** |
| Raw reads | 4160999 | 3392715 | 7957311 | 7131699 | 6665555 | 6032370 | 5312840 | 7703139 | 7439589 |
| Classified reads | 4067466 | 3346293 | 7898987 | 7079009 | 6631306 | 6014587 | 5267526 | 7641373 | 7389992 |
| Unclassified reads | 93533 | 46422 | 58324 | 52690 | 34249 | 17783 | 45314 | 61766 | 49597 |
| Eukaryota | | | | | | | | | |
| *Homo sapiens* | 3956481 | 3281446 | 6717900 | 5788246 | 6355657 | 5911381 | 5033556 | 7609726 | 7306022 |
| Bacteria | 79611 | 53624 | 1155868 | 1260227 | 259648 | 88842 | 210243 | 20479 | 71905 |
| *Pseudomonas* | 75257 | 50266 | 1153468 | 1257109 | 257941 | 88234 | 208373 | 17343 | 68137 |
| *Klebsiella* | 1664 | 2059 | <1% reads | <1% reads | <1% reads | <1% reads | <1% reads | 1719 | 3432 |
| *Citrobacter* | <1% reads | 906 | <1% reads | <1% reads | <1% reads | <1% reads | <1% reads | <1% reads | <1% reads |
| *Staphylococcus* | <1% reads | <1% reads | <1% reads | <1% reads | <1% reads | <1% reads | <1% reads | 466 | <1% reads |
| Other bacteria[1] | 2690 | 393 | 2400 | 3118 | 1707 | 608 | 1870 | 951 | 336 |
| Viruses | 3330 | 3513 | 859 | 12579 | 3526 | 4252 | 8391 | 793 | 2627 |
| *Pakpunavirus* | 161 | 683 | 201 | 12406 | 3475 | 613 | 1988 | 334 | 1202 |
| *Pbunavirus* | 272 | 480 | 126 | <1% reads | <1% reads | 3351 | 5710 | 366 | 973 |
| *Litunavirus* | 2811 | 2202 | 494 | <1% reads | <1% reads | 119 | 324 | 46 | 192 |
| *Phikzvirus* | <1% reads | <1% reads | <1% reads | <1% reads | <1% reads | 59 | 179 | 21 | 132 |
| *Tequatrovirus* | 60 | 123 | 28 | <1% reads | <1% reads | <1% reads | <1% reads | <1% reads | <1% reads |
| *Bruynoghevirus* | <1% reads | <1% reads | <1% reads | <1% reads | <1% reads | 47 | 87 | 14 | 80 |
| *Phikmvvirus* | <1% reads | <1% reads | <1% reads | <1% reads | <1% reads | 54 | <1% reads | 8 | 29 |
| Other viruses[1] | 26 | 25 | 10 | 173 | 51 | 9 | 103 | 4 | 19 |

[1] Includes all other genera with fewer than 1% classified reads.

**Table S4: Quantitative computed tomography (qCT) measurements of lung physiology changes from second-line treatment.**

| | Sample Date and Metric | | | | | | | |
|---|---|---|---|---|---|---|---|---|
| | **D-2** | | | | **D16** | | | |
| **Region** | AT (%) | VI (L) | VE (L) | E (%) | AT (%) | VI (L) | VE (L) | E (%) |
| Lungs | 76.59 | 4.87 | 3.89 | 4.02 | 72.12 | 5.24 | 4.05 | 7.46 |
| Right Lung | 79.58 | 3.09 | 2.53 | 4.19 | 75.41 | 3.18 | 2.51 | 7.44 |
| Left Lung | 71.42 | 1.78 | 1.37 | 3.72 | 67.07 | 2.06 | 1.54 | 7.49 |
| Right Upper Lobe | 87.53 | 0.65 | 0.55 | 11.06 | 85.61 | 0.53 | 0.52 | 19.87 |
| Right Lower Lobe | 75.68 | 2.1 | 1.55 | 1.25 | 70.62 | 2.15 | 1.55 | 3.45 |
| Left Upper Lobe | 87.56 | 0.63 | 0.54 | 8.54 | 83.71 | 0.63 | 0.59 | 15.47 |
| Left Lower Lobe | 61.37 | 1.07 | 0.76 | 0.4 | 59.15 | 1.38 | 0.88 | 3.42 |

Abbreviations: AT, air trapping; VI, volume inspiratory; VE, volume expiratory; E, emphysema.

**Table S5: Patient's clinical metrics from pulmonary and hematology tests at UCSD Medical Center.**

| | Treatment Period and Sample Date | | | | | | | | | | | | | | | |
|---|---|---|---|---|---|---|---|---|---|---|---|---|---|---|---|---|
| | Pre-treatment (baseline) | | | | Colistin/ceftazidime | Antibiotic holiday | | Ciprofloxacin | Ciprofloxacin/phage | | | | Post-treatment | | | |
| **Health metric** | D-202 | D-132 | D-111 | D-69 | D-13 | D-8 | D-5 | D-1 | D1 | D7 | D10 | D15 | D29 | D44 | D78 | D120 |
| $FEV_1$ (L) | 1.52 | 1.4 | 1.5 | 1.55 | 0.92 | 1.24 | 1.54 | 1.66 | 1.74 | 1.74 | 1.7 | 1.44 | 1.32 | 1.38 | 1.3 | 1.22 |
| FVC (L) | 3.13 | 2.82 | 2.98 | 3.06 | 1.69 | 2.44 | 2.75 | 3.03 | 3.28 | 3.32 | 3.19 | 3.01 | 2.69 | 3 | 2.91 | 2.95 |
| FEV1/FVC (%) | 49 | 50 | 50 | 51 | 54 | 51 | 56 | 55 | 53 | 52 | 53 | 48 | 49 | 46 | 45 | 41 |
| White blood cells ($\mu L^{-1}$) | 5400 | 4200 | 4500 | 7400 | 8200 | 7700 | 7500 | 6700 | 7000 | 7900 | 6800 | 7400 | 6500 | 8500 | 7000 | 10450 |
| Platelets ($\mu L^{-1}$) | 213000 | 204000 | 225000 | 230000 | 263000 | 307000 | 270000 | 230000 | 216000 | 234000 | 201000 | 161000 | 200000 | 251000 | 303000 | 318000 |
| Creatinine (mg/dL) | 1.17 | 1.24 | 1.29 | 1.3 | 1.22 | 2.93 | 3.79 | 2.77 | 2.87 | 2.86 | 2.45 | 2.32 | 2.12 | 1.8 | 1.54 | 1.5 |
| MMEF (L/s) | 0.47 | 0.49 | 0.45 | 0.5 | 0.35 | 0.45 | 0.63 | 0.6 | 0.62 | 0.58 | 0.65 | 0.44 | 0.48 | 0.48 | 0.39 | 0.33 |
| Percent predicted $FEV_1$ ($ppFEV_1$ %) | 44 | 41 | 43 | 45 | 27 | 36 | 45 | 48 | 50 | 51 | 49 | 42 | 38 | 40 | 38 | 41 |
| Percent predicted FVC (ppFVC %) | 68 | 61 | 65 | 67 | 37 | 53 | 60 | 66 | 72 | 72 | 70 | 66 | 59 | 65 | 63 | 64 |

Abbreviations: $FEV_1$, forced expiratory volume in 1 second; FVC, forced vital capacity; MMEF, maximal mid-expiratory flow.

**Table S6: Absolute principal component loadings for each variable.**

| **Health metric** | **Principal components (PC)** | | | | | | | | |
|---|---|---|---|---|---|---|---|---|---|
| | PC1 | PC2 | PC3 | PC4 | PC5 | PC6 | PC7 | PC8 | PC9 |
| Forced vital capacity (FVC) | 2.8795 | **3.6583** | 0.1448 | 0.0828 | 0.0510 | 0.0051 | 0.2730 | 0.2564 | **0.7451** |
| Forced expiratory volume in 1 s ($FEV_1$) | **3.3580** | 0.0066 | 0.0216 | 0.0806 | 0.1708 | 0.1723 | 0.3035 | **0.7967** | 0.0739 |
| Creatinine | 1.6112 | 2.8005 | 0.3811 | 0.1303 | **0.7186** | 0.2114 | 0.0002 | 0.0105 | 0.0002 |
| Maximal mid expiratory flow (MMEF) | 3.3394 | 1.9538 | 0.0022 | 0.0104 | 0.0951 | **0.8464** | 0.0992 | 0.0184 | 0.0020 |
| $FEV_1$/FVC | 1.2024 | 2.0577 | 0.2446 | 0.0147 | 0.4406 | 0.3453 | 0.2803 | 0.3555 | 0.0603 |
| %predicted FVC | 2.9100 | 2.0035 | 0.1406 | 0.0523 | 0.0351 | 0.0139 | 0.3207 | 0.4092 | 0.6595 |
| %predicted $FEV_1$ | 3.2462 | 0.4015 | 0.1398 | 0.0898 | 0.2312 | 0.2943 | **0.7946** | 0.0714 | 0.0163 |
| White blood cell counts | 1.1523 | 0.0436 | **0.6867** | 0.5670 | 0.4184 | 0.0228 | 0.0829 | 0.0113 | 0.0160 |
| Platelet counts | 1.7316 | 0.8370 | 0.5124 | **0.7982** | 0.1373 | 0.0510 | 0.0619 | 0.0064 | 0.0146 |
| Proportion of variance | 0.4348 | 0.2792 | 0.0461 | 0.0370 | 0.0466 | 0.0398 | 0.0450 | 0.0393 | 0.0322 |
| Cumulative proportion of variance | 0.4348 | 0.7139 | 0.7601 | 0.7971 | 0.8437 | 0.8836 | 0.9285 | 0.9678 | 1.0000 |

[1] **Bolded** components indicate the major variable loadings for a given component.

**Table S7: Mapping statistics of treatment phage genomes.**

| Sample | Phage | Genome coverage[1] | Plus reads | Minus reads |
|---|---|---|---|---|
| D-321 | P1A | 32.25% | 163 | 161 |
| | PYO2 | 99.82% | 2832 | 2836 |
| D-111 | P1A | 29.28% | 688 | 686 |
| | PYO2 | 99.77% | 2243 | 2242 |
| D-69 | P1A | 29.48% | 170 | 170 |
| | PYO2 | 59.85% | 425 | 427 |
| D3 | P1A | 100.00% | 12463 | 12464 |
| | PYO2 | 15.76% | 59 | 59 |
| D7 | P1A | 99.66% | 3501 | 3501 |
| | PYO2 | 66.90% | 24 | 24 |
| D9 | P1A | 76.61% | 606 | 611 |
| | PYO2 | 30.75% | 122 | 122 |
| D11 | P1A | 98.98% | 1991 | 1982 |
| | PYO2 | 62.08% | 322 | 326 |
| D12 | P1A | 53.23% | 337 | 335 |
| | PYO2 | 13.38% | 47 | 46 |
| D29 | P1A | 92.73% | 1209 | 1209 |
| | PYO2 | 40.35% | 196 | 197 |

[1] Coverage based on mapping to genome lengths P1A = 93,237 and PYO2 = 72,697.
